# Incidence Rates of Pediatric Central Nervous Tumors Display No Geographical Variation

**DOI:** 10.1101/2021.09.27.21264205

**Authors:** Dayton Grogan, Vamsi Reddy, Christopher Banerjee, Jennifer Waller, Theodore Johnson, Ian Heger

## Abstract

**Background:** Central nervous system (CNS) tumors affect over 4,600 children throughout the United States each year. Despite recent trends of increasing incidence of pediatric CNS tumors, the understanding of variations in their incidence between different geographical regions remains incomplete.

**Methods:** Data used in this study was obtained from the Surveillance, Epidemiology, and End Results (SEER) Program. The SEER database and its built-in operation software was used to generate state-specific incidence data for newly diagnosed CNS tumor diagnoses in children ages 0-19 for the years 2001-2014. Results were organized by tumor type and individual states were clustered into nine geographical regions as defined by the United States Census Bureau.

**Results:** Statistically significant differences were found in the regional incidence of astrocytoma, primitive neuroectodermal tumor (PNET), and the category of unspecified intracranial and intraspinal neoplasms. However, the magnitude of the difference in incidence (ΔI) between specific regions was small, on the order of 0.1 to 0.6 per 100,000 population, representing a nominal 0.05-fold to 0.79-fold change in incidence (ΔI/incidence for comparator region) for astrocytoma and for the category of unspecified intracranial and intraspinal neoplasms, and a larger 3.25-fold to 3.75-fold change in incidence for PNET.

**Conclusions:** Differences in incidence between geographical regions for certain CNS tumor types met the bar for statistical significance. However, these differences are unlikely to be clinically meaningful due to the small effect size.

## Introduction

Tumors of the central nervous system (CNS) present significant morbidity and mortality for affected children in the United States. According to the Central Brain Tumor Registry of the United States (CBTRUS), the incidence of primary malignant and nonmalignant CNS tumors in children and adolescents 0-19 years of age is 5.6 cases per 100,000^1^. Collectively, CNS tumors are among the most common childhood malignancies, second only to hematologic malignancies, and are the most common form of pediatric solid organ tumor^2^. CNS tumors also represent 20% of all childhood malignancies and are the leading cause of cancer-related deaths in children ages 0-14^3^. Furthermore, according to recent CDC reports, childhood cancer deaths are in decline except for pediatric CNS tumors^3^. Therefore, understanding environmental influences that may impact the risk for developing childhood CNS cancer is critical to devising prevention strategies.

Over the past two decades, the overall reported incidence of pediatric CNS tumors has increased^4-10^. However, it is possible that some of this increase may be artifactual, as improvements in detection capabilities have led to an increase in documented CNS tumor cases^11-16^. Moreover, changes in histological subtyping of tumors, alterations of registry code assignments, and recording of benign tumors in cancer registries have also impacted overall incidence reports. While such factors are unlikely to account for the entire increment in pediatric CNS tumor incidence, the effects of these systemic factors may complicate the study of environmental exposures that may genuinely impact tumorigenesis. To begin elucidating the contribution of such factors, a publically available cancer registry was used to determine whether incidence rates of pediatric CNS tumors vary by geographical location.

## Methods

### Data Acquisition

All data utilized in this study was obtained from the Surveillance, Epidemiology, and End Results (SEER) Program. This program supported by the National Cancer Institute provides researchers with analytical tools and methodological capability for analysis of population-based statistics in the United States. The SEER Program comprises a database of cancer incidence and survival information collected from population-based cancer registries accounting for approximately 34.6% of the U.S. population^17^. Demographically, the SEER program includes data from a significant proportion of cancer patients in the following racial/ethnic populations: Caucasian (31.9%), African American (30%), Hispanic (44%), American Indian/Alaska Native (49.3%), Asians (57.5%), and Hawaiian/Pacific Islander (68.5%). The SEER database and its built-in operation software was used to generate state-specific incidence data for new CNS tumors diagnoses in children ages 0-19 for the years 2001-2014. For a particular tumor type category, data was withheld if there was not a minimum of 16 cases per year, in order to protect against identification of patients with rare tumor types.

### Data Organization

For analyses of differential incidence rates, states were organized into nine geographical regions as defined by the United States Census Bureau (**Table 1**). Incidence values for individual states within each geographical region were averaged based on data from the SEER program, and the mean incidence (with 95% confidence interval) were calculated for each region. Incidence data was subdivided according to CNS tumor type, using the predetermined subgrouping system used by the SEER database.

**Table 1:** Regions used in this study with affiliated states.

| **Region** | **Associated States** |
| --- | --- |
| **New England** | Connecticut, Maine, Massachusetts, New Hampshire, Rhode Island, Vermont |
| **Mid-Atlantic** | New Jersey, New York, Pennsylvania |
| **South Atlantic** | Delaware, Florida, Georgia, Maryland, North Carolina, South Carolina, Virginia, District of Columbia |
| **East North Central** | Illiois, Indiana, Michigan, Ohio, Wisconsin |
| **East South Central** | Alabama, Kentcky, Mississippi, Tennessee |
| **West North Central** | Iowa, Kansas, Minnesota, Missouri, Nebraska, North Dakota, South Dakota |
| **West South Central** | Arkansas, Louisiana, Oklahoma, Texas |
| **Mountain** | Arizona, Colorado, Idaho, Montana, Nevada, New Mexico, Utah, Wyoming |
| **Pacific** | Alaska, California, Hawaii, Oregon, Washington |

### Statistical Analysis

All statistical analysis was performed using SAS 9.4 and statistical significance was assessed using an alpha level of 0.05. Tumor incidence rates/100,000 within region were determined for each tumor type and sub-type from the SEER program database. To examine differences in incident rates between regions for each tumor type and sub-type, a generalized linear model was used with a negative binomial distribution for the number of incident cases, a log link, and an offset parameter of the log of the pediatric population at risk. A Tukey-Kramer multiple comparison test was used to examine post hoc pairwise differences between regions.

## Results

Combined incidence for all pediatric CNS tumors from 2001-2014 were analyzed and plotted onto a map of the United States in order to examine apparent, overarching trends (**Figure 1**). The incidence data was then subdivided into specific CNS tumor types, and heat maps were generated for each tumor subtype (data not shown). Superficial regional differences were indicated by the heat maps for malignant tumors, suggesting an overall higher incidence occurring in the New England and Mid-Atlantic regions.

**Figure 1.**
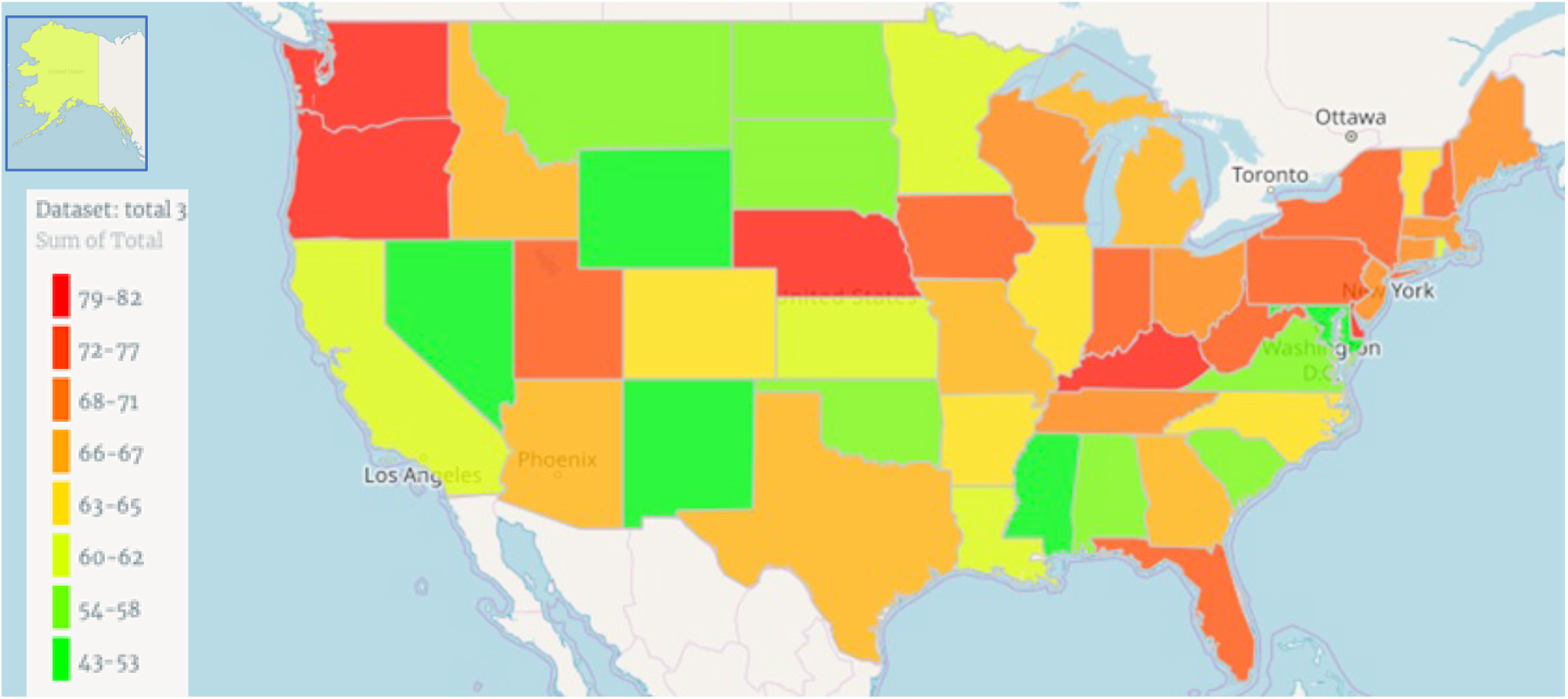
Heat map demonstrating relative trends in regional differences of total incidence reports for pediatric CNS tumors from 2001-2014. Values were determined by dividing the sum of all reported incidence values for all pediatric CNS tumors by the pediatric population (0-19) of each state per 100,000 per year.

Further analysis was required to demonstrate which regions may have statistically significant differences in incidence rates of specific CNS tumor types. ANOVA and subsequent pairwise analyses were performed to examine these differences, as summarized in **Table 2**. This analysis revealed statistically significant differences in the regional incidence of astrocytoma (p=0.0024), primitive neuroectodermal tumor (PNET, p=0.0153), and unspecified intracranial and intraspinal neoplasms (p<0.0001). However, the magnitude of the difference in incidence (ΔI) between specific regions was small, on the order of 0.1 to 0.6 per 100,000 population, representing a nominal 0.05-fold to 0.79-fold change in incidence (ΔI/incidence for comparator region) for astrocytoma and for unspecified intracranial and intraspinal neoplasms, and a larger 3.25-fold to 3.75-fold change in incidence for PNET.

**Table 2:** Incidence rates per 100,000 (95% Confidence Interval) by Region within Tumor Type or Subtype and Generalized Linear Model Test for Differences Between Regions.

| Tumor Type/Subtype | East North-Central | East South-Central | Mid-Atlantic | Mountain | New England | Pacific | South Atlantic | West North-Central | West South-Central | $\chi^2$ | p-value |
| --- | --- | --- | --- | --- | --- | --- | --- | --- | --- | --- | --- |
| III CNS and Miscellaneous intracranial and intraspinal neoplasms | 5.4<br>(5.00, 5.8)0 | 5.5<br>(4.80, 6.20) | 6.4<br>(5.90, 6.90) | 5.3<br>(4.70, 5.90) | 5.6<br>(4.80, 6.40) | 5.3<br>(4.90, 5.70) | 5.6<br>(5.20, 6.00) | 5.6<br>(5.00, 6.30) | 5.7<br>(5.30, 6.30) | 14.87 | 0.0616 |
| III (a) Ependymomas and choroid plexus tumor | 0.42<br>(0.36, 0.50) | 0.41<br>(0.32, 0.54) | 0.47<br>(0.39, 0.55) | 0.46<br>(0.37, 0.58) | 0.48<br>(0.36, 0.63) | 0.44<br>(0.38, 0.51) | 0.42<br>(0.36, 0.48) | 0.46<br>(0.37, 0.58) | 0.46<br>(0.39, 0.54) | 2.28 | 0.971 |
| III (a.1) Ependymomas | 0.32<br>(0.26, 0.39) | 0.32<br>(0.24, 0.44) | 0.36<br>(0.29, 0.44) | 0.36<br>(0.28, 0.47) | 0.35<br>(0.25, 0.50) | 0.32<br>(0.27, 0.39) | 0.32<br>(0.27, 0.38) | 0.35<br>(0.26, 0.46) | 0.37<br>(0.30, 0.45) | 2.36 | 0.9679 |
| III (a.2) Choroid plexus tumor | 0.11<br>(0.07, 0.16) | 0.06<br>(0.02, 0.14) | 0.11<br>(0.07, 0.17) | 0.08<br>(0.04, 0.15) | 0.08<br>(0.03, 0.19) | 0.1<br>(0.07, 0.15) | 0.09<br>(0.06, 0.13) | 0.11<br>(0.06, 0.20) | 0.07<br>(0.04, 0.12) | 4.47 | 0.8125 |
| III (b) Astrocytoma | 2<br>(1.90, 2.20) | 2<br>(1.80, 2.30) | 2.4<br>(2.20, 2.60) | 1.9<br>(1.70, 2.20) | 2.1<br>(1.80, 2.50) | 1.8<br>(1.70, 2.00) | 2<br>(1.90, 2.20) | 2.2<br>(2.00, 2.50) | 2<br>(1.80, 2.20) | 23.84 | 0.0024 |
| III (c) Intracranial and intraspinal embryonal tumors | 0.74<br>(0.68, 0.80) | 0.68<br>(0.60, 0.78) | 0.71<br>(0.65, 0.78) | 0.76<br>(0.68, 0.85) | 0.68<br>(0.58, 0.79) | 0.77<br>(0.72, 0.83) | 0.73<br>(0.68, 0.78) | 0.75<br>(0.66, 0.84) | 0.77<br>(0.70, 0.83) | 5.93 | 0.6552 |
| III (c.1) Medulloblastomas | 0.5<br>(0.44, 0.56) | 0.45<br>(0.36, 0.55) | 0.43<br>(0.37, 0.50) | 0.48<br>(0.40, 0.58) | 0.42<br>(0.33, 0.54) | 0.48<br>(0.43, 0.55) | 0.46<br>(0.41, 0.52) | 0.53<br>(0.44, 0.64) | 0.52<br>(0.45, 0.59) | 6.71 | 0.5687 |
| III (c.2) PNET | 0.16<br>(0.11, 0.22) | 0.11<br>(0.06, 0.21) | 0.17<br>(0.12, 0.24) | 0.08<br>(0.04, 0.16) | 0.04<br>(0.01, 0.13) | 0.19<br>(0.14, 0.25) | 0.14<br>(0.10, 0.19) | 0.04<br>(0.02, 0.11) | 0.12<br>(0.08, 0.19) | 18.92 | 0.0153 |
| III (c.3) Medulloepithelioma | -- | -- | -- | -- | -- | -- | -- | -- | -- | -- | -- |
| III (c.4) Atypical teratoid/rhabdoid tumor | 0.07<br>(0.04, 0.12) | 0.09<br>(0.04, 0.18) | 0.09<br>(0.06, 0.15) | 0.08<br>(0.04, 0.16) | 0.05<br>(0.01, 0.15) | 0.09<br>(0.06, 0.13) | 0.1<br>(0.07, 0.15) | 0.06<br>(0.02, 0.13) | 0.11<br>(0.07, 0.17) | 4.33 | 0.8263 |
| III (d) Other gliomas | 0.7<br>(0.57, 0.84) | 0.66<br>(0.48, 0.91) | 0.82<br>(0.67, 1.00) | 0.56<br>(0.41, 0.76) | 0.75<br>(0.53, 1.10) | 0.62<br>(0.51, 0.75) | 0.69<br>(0.57, 0.82) | 0.62<br>(0.46, 0.85) | 0.76<br>(0.62, 0.93) | 7.51 | 0.4831 |
| III (d.1) Oligodendrogliomas | 0.08<br>(0.05, 0.13) | 0.03<br>(0.01, 0.11) | 0.1<br>(0.06, 0.16) | 0.03<br>(0.01, 0.10) | 0.08<br>(0.03, 0.20) | 0.08<br>(0.05, 0.12) | 0.07<br>(0.04, 0.12) | 0.07<br>(0.03, 0.15) | 0.04<br>(0.02, 0.08) | 7.85 | 0.4483 |
| III (d.2) Mixed and unspecified gliomas | 0.6<br>(0.48, 0.73) | 0.59<br>(0.42, 0.83) | 0.7<br>(0.57, 0.87) | 0.46<br>(0.33, 0.65) | 0.65<br>(0.45, 0.94) | 0.53<br>(0.42, 0.65) | 0.58<br>(0.48, 0.71) | 0.51<br>(0.36, 0.72) | 0.69<br>(0.56, 0.85) | 8.53 | 0.3839 |
| III (d.3) Neuroepithelial glial tumors of uncertain origin | -- | -- | -- | -- | -- | -- | -- | -- | -- | -- | -- |
| III (e) Other specified intracranial/intraspinal neoplasms | 1.4<br>(1.20, 1.60) | 1.4<br>(1.10, 1.80) | 1.8<br>(1.60, 2.00) | 1.3<br>(1.10, 1.60) | 1.3<br>(1.00, 1.80) | 1.5<br>(1.30, 1.70) | 1.5<br>(1.30, 1.70) | 1.4<br>(1.10, 1.70) | 1.4<br>(1.20, 1.60) | 10.65 | 0.2222 |
| III (e.1) Pituitary adenomas and carcinomas | 0.52<br>(0.39, 0.69) | 0.53<br>(0.34, 0.84) | 0.87<br>(0.68, 1.10) | 0.56<br>(0.38, 0.84) | 0.48<br>(0.28, 0.84) | 0.71<br>(0.56, 0.90) | 0.6<br>(0.47, 0.76) | 0.56<br>(0.37, 0.85) | 0.62<br>(0.46, 0.82) | 11.1 | 0.1648 |
| III (e.2) Tumors of sellar region (craniopharyngiomas) | 0.18<br>(0.14, 0.24) | 0.23<br>(0.16, 0.35) | 0.24<br>(0.18, 0.31) | 0.15<br>(0.10, 0.24) | 0.12<br>(0.06, 0.23) | 0.2<br>(0.15, 0.26) | 0.2<br>(0.16, 0.26) | 0.13<br>(0.08, 0.22) | 0.22<br>(0.17, 0.29) | 9.85 | 0.276 |
| III (e.3) Pineal parenchymal tumors | 0.05<br>(0.03, 0.08) | -- | 0.05<br>(0.03, 0.08) | -- | -- | 0.05<br>(0.03, 0.08) | 0.04<br>(0.03, 0.07) | -- | 0.04<br>(0.02, 0.07) | -- | -- |
| III (e.4) Neuronal and mixed neuronal-glial tumors | 0.47<br>(0.39, 0.58) | 0.42<br>(0.30, 0.59) | 0.45<br>(0.36, 0.56) | 0.35<br>(0.25, 0.49) | 0.46<br>(0.31, 0.67) | 0.37<br>(0.30, 0.46) | 0.42<br>(0.34, 0.51) | 0.41<br>(0.30, 0.57) | 0.37<br>(0.28, 0.47) | 5.64 | 0.6876 |
| III (e.5) Meningiomas | 0.16<br>(0.12, 0.22) | 0.19<br>(0.12, 0.29) | 0.17<br>(0.13, 0.24) | 0.1<br>(0.06, 0.18) | 0.09<br>(0.04, 0.20) | 0.14<br>(0.10, 0.19) | 0.15<br>(0.11, 0.20) | 0.16<br>(0.10, 0.25) | 0.12<br>(0.08, 0.18) | 6.64 | 0.5757 |
| III (f) Unspecified intracranial and intraspinal neoplasms | 0.13<br>(0.09, 0.2) | 0.2<br>(0.12, 0.34) | 0.21<br>(0.15, 0.30) | 0.13<br>(0.07, 0.23) | 0.11<br>(0.05, 0.26) | 0.12<br>(0.08, 0.18) | 0.21<br>(0.16, 0.28) | 0.08<br>(0.03, 0.17) | 0.38<br>(0.29, 0.50) | 41.91 | <0.0001 |

For astrocytoma, the East North-Central region had significantly higher incidence (2/100,000) than the Mountain (ΔI=0.1/100,000; 0.05-fold change; p<0.0001), New England (ΔI=0.1/100,000; 0.05-fold change; p=0.0002), and West North-Central (ΔI=0.2/100,000; 0.1-fold change; p=0.0008) regions. The East South-Central region had significantly lower incidence (2/100,000) than the Mid-Atlantic region (ΔI=0.4/100,000; 0.2-fold change; p=0.0009), but significantly higher incidence than the Mountain (ΔI=0.1/100,000; 0.05-fold change; p<0.0001) region. The Mid-Atlantic region had significantly higher incidence (2.4/100,000) than the Mountain (ΔI=0.5/100,000; 0.21-fold change; p<0.0001), New England (ΔI=0.3/100,000; 0.13-fold change; p<0.0001), Pacific (ΔI=0.6/100,000; 0.25-fold change; p=0.0009), South Atlantic (ΔI=0.4/100,000; 0.17-fold change; p=0.0242), West North-Central (ΔI=0.2/100,000; 0.08-fold change; p<0.0001), and West South-Central (ΔI=0.4/100,000; 0.17-fold change; p=0.0015). The Mountain region had significantly lower incidence (1.9/100,000) than the Pacific (ΔI=0.1/100,000; 0.05-fold change; p<0.0001), South Atlantic (ΔI=0.1/100,000; 0.05-fold change; p<0.0001), West North-Central (ΔI=0.3/100,000; 0.16-fold change; p=0.0103), and West South-Central (ΔI=0.1/100,000; 0.05-fold change; p<0.0001).

PNET tumors were significantly underrepresented in the West North Central region, which had significantly lower incidence (0.04/100,000) than the Mid-Atlantic region (ΔI=0.13/100,000; 3.25-fold change; p=0.0072) and the Pacific region (ΔI=0.15/100,000; 3.75-fold change; 0.0039).

Regarding unspecified intracranial and intraspinal neoplasms, the West South-Central region had significantly higher incidence (0.38/100,000) than the East North Central (ΔI=0.25/100,000; 0.66-fold change; p=0.0007), Mountain (ΔI=0.25/100,000; 0.66-fold change; p=0.0296), Pacific (ΔI=0.26/100,000; 0.68-fold change; p=0.0001), and West North Central (ΔI=0.3/100,000; 0.79-fold change; p=0.0071) regions.

## Discussion

Pediatric brain cancer is rare, and one of the pitfalls in examining the geographical distribution of pediatric CNS tumor cases is the overall low incidence compared to the incidence in adult populations.^1,3^ This makes it necessary to analyze a prolonged time period and aggregate state-level data into geographical regions, and makes sub-stratification more difficult for rare tumor types. In our study, spanning the time period between 2001 and 2014, the raw state-level data revealed a trend of higher total pediatric CNS tumor incidence in certain states within the New England and Mid-Atlantic regions, which is similar to results from recent CDC epidemiological studies^18^.

Aggregation of state-level data into multi-state geographical regions allowed for statistical analysis to compare incidence rates. No statistically significant difference in the total pediatric CNS tumor incidence was found between the geographical regions. When the incidence data was stratified according to CNS tumor type, regional differences emerged with statistical significance in the tumor categories of astrocytoma, PNET, and unspecified intracranial and intraspinal neoplasms. However, such statistical differences arise from relatively minor differences between regions. For example, the incidence of astrocytoma for the Mid-Atlantic is slightly higher than the West South Central region, but the actual difference in incidence is 0.4 patients per 100,000 children (**Table 2**), which represents a trivial 0.17-fold difference. Using census-recorded pediatric population data for the Mid-Atlantic region during 2014 (8,939,737) as reference, this would translate into a small absolute difference of approximately 35 patients per year between these two regions. In the PNET category, the West North Central region had incidence rates that were 3.25-fold less than the Mid-Atlantic region and 3.75-fold less than the Pacific region. However, the actual incidence rates were very low (0.04 to 0.15/100,000), thirteen to fifty times lower than the incidence for astrocytoma, which translated to exceedingly low differences in absolute case numbers. Thus, while statistically significant regional differences emerge with stratification of the incidence data by CNS tumor type, the differences are very small and unlikely to be clinically meaningful.

Limitations for this study are related to data collection methods. The low overall incidence of pediatric CNS tumors limits the degree of sub-stratification that can maintain meaningful statistical results. In addition, the data collection methods utilized in the SEER data platform make it difficult to correlate differences in incidence to environmental factors. For instance, it is possible that a patient entered into the registry may have recently moved to that state (region), having spent the majority of their life in another geographical region. To protect patient privacy, incidence data is not reported for tumor types with less than sixteen cases per year, so the current study is not applicable to very rare pediatric CNS tumor types. Finally, future analyses of clinical registries will be hampered by evolving changes in the World Health Organization (WHO) classification of CNS tumor types, and it may not be possible to re-classify patients already in the database using the newer diagnostic criteria which rely heavily upon genetic analyses of tumors^19,20^.

Numerous previous studies have found significant regional differences in non-CNS tumors such as pleural mesothelioma, prostate cancer, and hepatocellular carcinoma.^21-23^ Such findings in non-CNS tumors were further correlated with regional and environmental differences in demographic factors such as age, gender, age, ethnicity, diet, *etc*. We speculate that certain pediatric tumors may follow similar trends, especially when considering potential effects of certain maternal environmental exposures.^24^ However, in this study no clinically meaningful differences were found between geographical regions in the incidence of pediatric CNS tumors, either in aggregate or stratified by CNS tumor type.

## Data Availability

All data was obtained from the Surveillance, Epidemiology, and End Results (SEER) Program.

